# Multifaceted strategies for the control of COVID-19 outbreaks in long-term care facilities in Ontario, Canada

**DOI:** 10.1101/2020.12.04.20244194

**Authors:** Thomas N. Vilches, Shokoofeh Nourbakhsh, Kevin Zhang, Lyndon Juden-Kelly, Lauren E. Cipriano, Joanne M. Langley, Pratha Sah, Alison P. Galvani, Seyed M. Moghadas

## Abstract

The novel coronavirus disease 2019 (COVID-19) has caused severe outbreaks in Canadian long-term care facilities (LTCFs). In Canada, over 80% of COVID-19 deaths during the first pandemic wave occurred in LTCFs. We sought to evaluate the effect of mitigation measures in LTCFs including frequent testing of staff, and vaccination of staff and residents. We developed an agent-based transmission model and parameterized it with disease-specific estimates, temporal sensitivity of nasopharyngeal and saliva testing, results of vaccine efficacy trials, and data from initial COVID-19 outbreaks in LTCFs in Ontario, Canada. Characteristics of staff and residents, including contact patterns, were integrated into the model with age-dependent risk of hospitalization and death. Estimates of infection and outcomes were obtained and 95% credible intervals were generated using a bias-corrected and accelerated bootstrap method. Weekly routine testing of staff with 2-day turnaround time reduced infections among residents by at least 25.9% (95% CrI: 23.3% - 28.3%), compared to baseline measures of mask-wearing, symptom screening, and staff cohorting alone. A similar reduction of hospitalizations and deaths was achieved in residents. Vaccination averted 2-4 times more infections in both staff and residents as compared to routine testing, and markedly reduced hospitalizations and deaths among residents by 95.9% (95% CrI: 95.4% - 96.3%) and 95.8% (95% CrI: 95.5% - 96.1%), respectively, over 200 days from the start of vaccination. Vaccination could have a substantial impact on mitigating disease burden among residents, but may not eliminate the need for other measures before population-level control of COVID-19 is achieved.

## Introduction

The novel coronavirus disease 2019 (COVID-19) has led to severe outbreaks in Canadian long-term care facilities (LTCFs). LTCF residents are particularly vulnerable to COVID-19 due to a high prevalence of comorbid conditions (Ontario Long Term Care Association, 2019) and their advanced age. Since the start of the COVID-19 pandemic, a number of strategies have been implemented to prevent infection and disease transmission in LTCFs, including non-pharmacological measures such as isolation, visitor restrictions, hand hygiene, and mask-wearing (Centers for Disease Control and Prevention, 2020; J. Wang et al., 2020). While COVID-19 mitigation measures have had a significant impact on reducing transmission on a population level (Zhang et al., 2020), control of outbreaks in LTCFs has proved to be challenging, in part due to silent transmission from infected asymptomatic or presymptomatic visitors and staff (Ladhani et al., 2020; Ouslander and Grabowski, 2020).

In Canada, over 80% of reported COVID-19 deaths during the first wave were among residents of LTCFs (Barnett and Grabowski, 2020; Canadian Institute for Health Information, 2020; Hsu et al., 2020; Webster, 2021). While advanced age and comorbid medical conditions are risk factors for a more severe course of disease among residents (Garnier□Crussard et al., 2020; B. Wang et al., 2020; Yang et al., 2020), recent studies have highlighted the inadequacy of the systemic response to COVID-19 in Canadian LTCFs (Brown et al., 2020; Liu et al., 2020; Stall et al., 2020). Shortages of staff and personal protective equipment (PPE), limited testing capacity with reliance on symptom-based screening, and inadequate space to implement efficient cohorting measures appear to have contributed to the extraordinary disease toll in these settings (Liu et al., 2020). Results from phase III clinical trials of COVID-19 vaccines, as well as their effectiveness in mass immunization campaigns indicate that prioritizing residents and staff for vaccination could prevent LTCF outbreaks (Baden et al., 2020; Dagan et al., 2021; Lipsitch and Kahn, 2021; Polack et al., 2020). However, whether vaccination alone is sufficient to protect these vulnerable populations from COVID-19 remains undetermined. Furthermore, the effectiveness of infection control measures in LTCFs depend on a number of factors such as the contact network between residents and staff, in addition to disease characteristics, such as the presence of symptoms and severity of illness at the individual level.

We sought to investigate the impact of various COVID-19 mitigation measures in LTCFs by developing an agent-based model of disease transmission dynamics. We parameterized the model with disease-specific estimates and data from initial outbreaks in LTCFs in the province of Ontario, Canada (Public Health Ontario, 2020). We also used movement and contact network data collected from the largest veterans care facility in Canada, located at Sunnybrook Health Sciences Centre (Champredon et al., 2018; Najafi et al., 2017). We evaluated the effect of case isolation, mask-wearing, cohorting and routine testing of staff in the absence of vaccination. We then expanded the model to include vaccination and evaluate the need for other interventions.

## Methods

### Ethics statement

The study was approved by the York University Ethics Review Board (Project: 2020-269). The study was categorized as minimal risk, with no requirement for individual consent or participation.

### Model structure and population

We developed an agent-based simulation model of COVID-19 transmission dynamics in a LTCF with resident and staff populations (Appendix, Figure A1). The model structure was informed by population demographics of LTCFs in Ontario, Canada, including age (Ontario Long Term Care Association, 2019; Statistics Canada, 2020), staff-to-resident ratio, distribution of rooms and occupancy (Stall et al., 2020). The interactions within and between staff and resident populations were parameterized using the distributions derived from close-range movement and contact network data collected through wearable sociometric tags in a Canadian LTCF (Najafi et al., 2017). The model included three working shifts of morning, evening, and night, each covering 8 hours of daily interactions.

Based on demographic data from the Ontario Long Term Care Association (Ontario Long Term Care Association, 2019), the resident population included individuals of age 50-64 (6.6%), 65-74 (11.4%), 75-84 (27.3%), 85-94 (43.9%), and 95+ (10.8%). A total of 120 residents (i.e., the average size of a LTCF) were included in the model and assigned to 84 rooms, with a distribution corresponding to 48 single and 36 double occupancy rooms.

The model considered a daily staff population of 68 individuals, aged 20-64 years of age (Statistics Canada, 2020), which included direct care providers (i.e., personal support workers, nurses), dietary staff and housekeeping personnel. The distribution of staff by classification and staff-to-resident ratio for each daily shift were informed by correspondence with the management teams of 10 LTFCs affected by COVID-19 outbreaks in Ontario. The overall number of staff varied in the model, depending on substitution that occurred for identified staff with infection.

Daily contacts among residents were sampled from a previously inferred distribution with a mean of 6.8 contacts per resident per day (Champredon et al., 2018; Najafi et al., 2017). Daily numbers of contacts varied from 6 to 9 between residents and direct care providers (nurses and personal support workers). Due to outbreak measures implemented in LTCFs, the model was parameterized with limited contacts between residents and other service staff based on correspondence with the management teams of LTFCs (Appendix, Table A1). Given visiting restrictions during outbreaks, we did not include visitors in the model. Contacts among staff were distributed in each shift depending on the responsibility of staff in the LTCF (Appendix, Table A1).

### Disease dynamics

We encapsulated the natural history of COVID-19 with epidemiological statuses as susceptible; latently infected (not yet infectious); asymptomatic (and infectious); pre-symptomatic (and infectious); symptomatic with either mild or severe/critical illness; recovered; and dead (Figure 1). We assumed that infection was introduced into the LTCF through infected staff during the silent asymptomatic or pre-symptomatic stages of disease. A population point-prevalence in the range of 0.05%-0.1% was considered for infection of staff outside the LTCF prior to the start of each shift (Defence Research and Development Canada, 2020). Staff with symptomatic COVID-19, whether infected outside or during the daily shifts in the LTCF, were screened and removed from the model simulations for a minimum of 14 days (and until complete recovery). Substitute staff were considered in the model to assume responsibilities of the infected staff during their isolation period.

**Figure 1.**
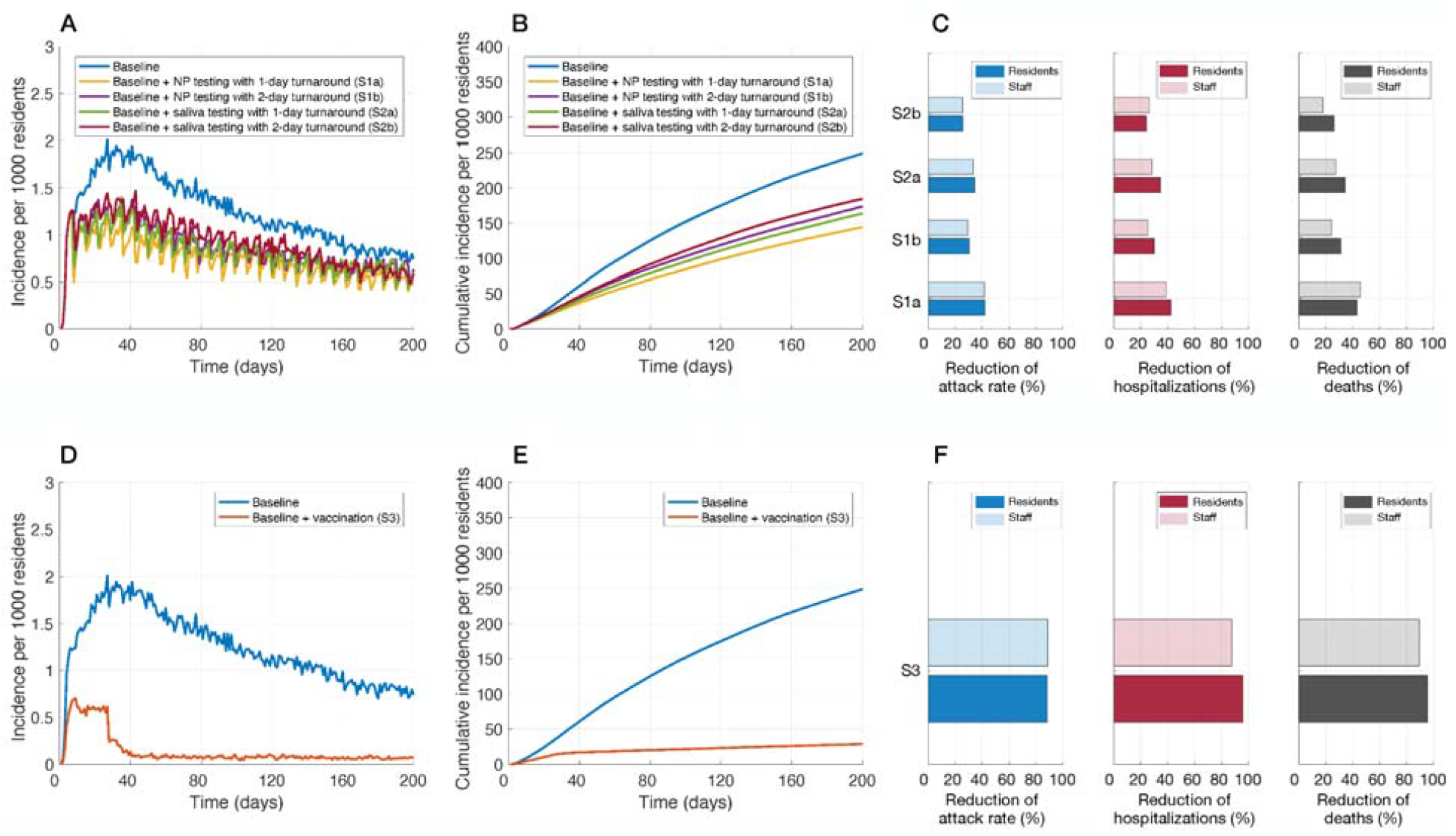
Incidence of infection per 1000 residents (A,D), cumulative infections per 1000 residents (B,E), and relative reduction of cumulative infections (attack rate), hospitalizations, and deaths with routine testing of staff (C) and vaccination of residents and staff (F), over a 200 time period of simulations. Intervention scenarios (in addition to baseline control measures) included: weekly routine testing of staff with NP sampling and 1-day (S1a) and 2-day (S1b) turnaround; saliva sampling and 1-day (S2a) and 2-day (S2b) turnaround; and vaccination (S3).

Disease transmission inside the LTCF was implemented probabilistically for contacts between susceptible and infectious individuals in asymptomatic, pre-symptomatic, or symptomatic stages of the disease. Infected individuals started in the latent (non-infectious) stage, and then proceeded to a silent infectious stage (i.e., either asymptomatic or pre-symptomatic). A proportion of infected individuals remained asymptomatic until recovery (Buitrago-Garcia et al., 2020; DeBiasi and Delaney, 2020; X. Li et al., 2020; Poline et al., 2020), with an infectious period that was sampled from a Gamma distribution with a mean of 5 days (Gatto et al., 2020; R. Li et al., 2020). Others developed symptoms following a pre-symptomatic stage as part of the incubation period. The incubation and pre-symptomatic periods were sampled from Log-Normal and Gamma distributions with mean values of 5.2 and 2.3 days, respectively (He et al., 2020; Lauer et al., 2020). The infectious period post-symptom onset was also sampled from a Gamma distribution with a mean of 3.2 days (R. Li et al., 2020). Symptomatic cases had an age-dependent probability of developing mild or severe/critical illness. We assumed that recovery from a primary infection provided adequate immunity for the remainder of the simulation, preventing re-infection. Compared to the probability of transmission during the pre-symptomatic stage, the relative risks were 0.26, 0.44, and 0.89 for the asymptomatic, mild symptomatic, and severe symptomatic stages, respectively (Ferretti et al., 2020; Moghadas et al., 2020a; Sayampanathan et al., 2021).

### Infection outcomes

Residents with symptomatic disease and their roommates, based on outbreak guidelines, were immediately isolated upon symptom onset within the LTCF (Ministry of Health, 2020). Contacts of isolated cases were limited to only direct healthcare providers. A proportion of symptomatic residents who developed severe illness were transferred and hospitalized, and therefore excluded from the dynamics within the LTCF until their return upon recovery. For those who were hospitalized, the time from symptom onset to admission was sampled in the range of 2-5 days (Moghadas et al., 2020b; Shoukat et al., 2020). The length of hospital stay was sampled from a Gamma distribution with mean of 12.4 days (Sanche et al., 2020). We parameterized the model with age-specific hospitalization rates of LTCF residents from January to June 2020 (Table 1). The case fatality rate among residents was age-dependent and based on data reported by Public Health Ontario (Table 1) (Public Health Ontario, 2020).

**Table 1.**
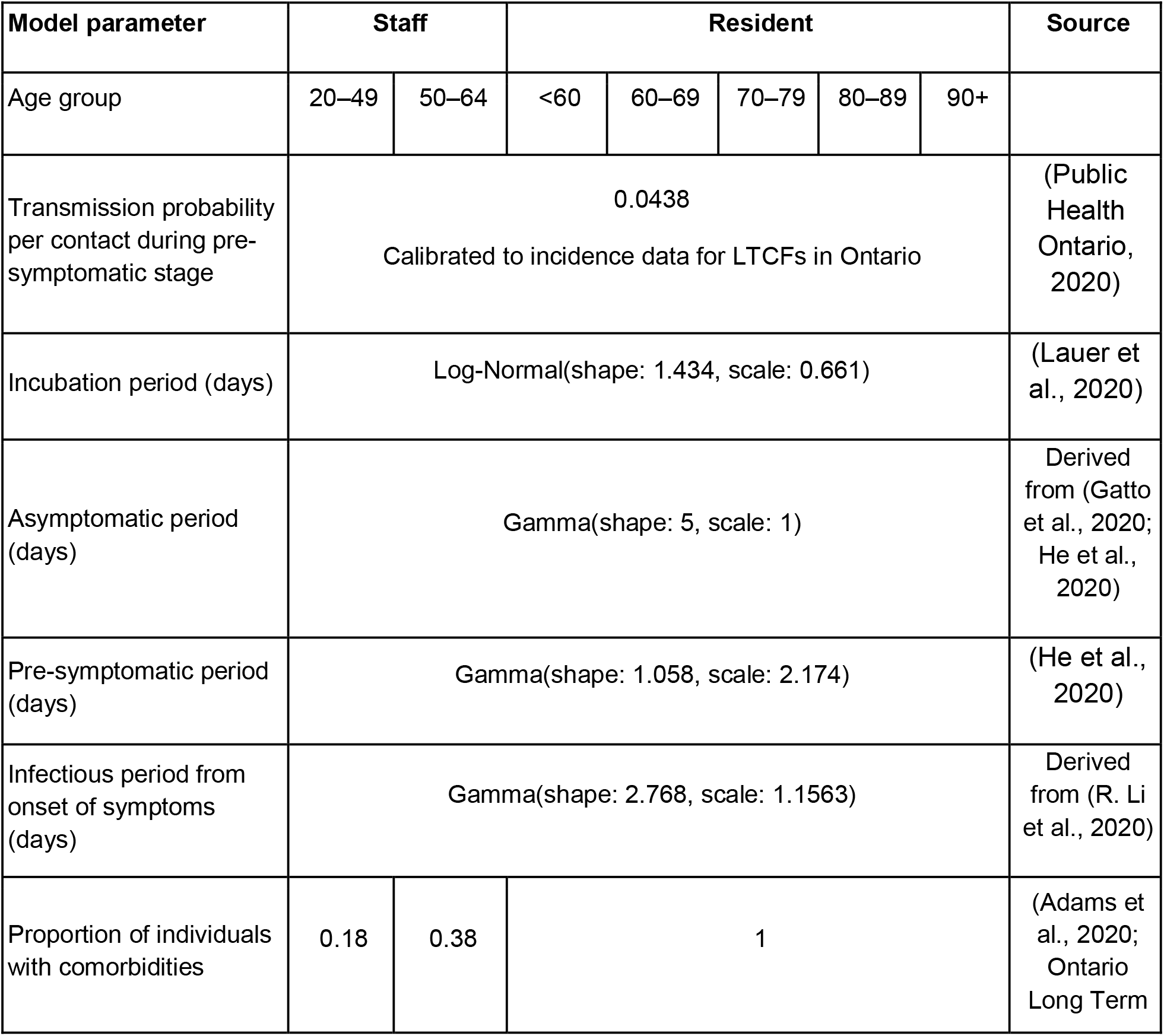
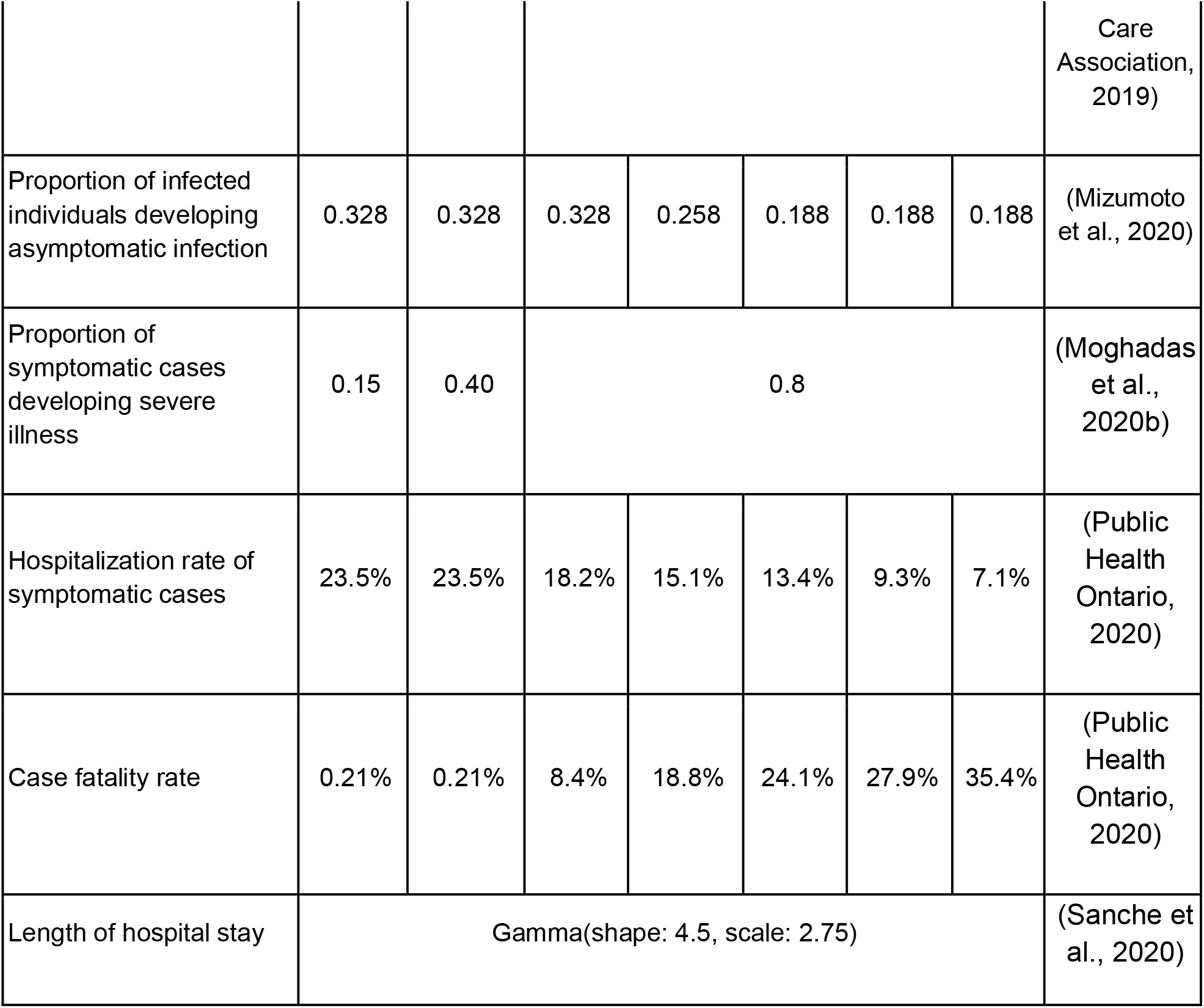
Model parameters and their estimated value.

### Interventions

The baseline scenario of control measures included: (i) isolation of symptomatic residents with hospitalization of a proportion who developed severe illness; (ii) screening of staff for symptomatic illness followed by isolation; (iii) cohorting of care providers; and (iv) mask-wearing by all staff. We assumed that all staff wore surgical masks during their shift, but switched to an N95 respirator when caring for isolated residents. A recent meta-analysis indicates a 67% (95% CI: 39% − 83%) risk reduction in respiratory infections when surgical masks are used (Chu et al., 2020). Thus, the transmission probability per contact was reduced by a factor (1-eff_sur_) for staff-resident, and (1-eff_sur_)^2^ for staff-staff interactions. When an N95 was used by staff caring for isolated residents, the probability of transmission was reduced by (1-eff_N95_) per contact, with an efficacy eff_N95_ = 0.95 (Chu et al., 2020). For staff cohorting, we assigned each healthcare provider to a specific group of residents. Personal support workers only interacted with a predetermined group of 9, 9, and 20 residents during the three daily shifts of morning, evening, and night. Similarly, nurses interacted with predetermined groups of 30, 30, and 60 residents during the corresponding shifts. Given these baseline interventions, we compared two additional measures: routine testing of all staff and vaccination of staff and residents.

#### Routine testing

To prevent disease importation and transmission during the silent asymptomatic or pre-symptomatic stages of infection, we implemented routine nasopharyngeal (NP) and saliva PCR testing of staff with a frequency of 7 days (Iacobucci, 2020). The probability of case detection at the time of testing post-infection was determined by the temporal diagnostic sensitivity (Zhang et al., 2021) inferred from fitting a sensitivity function to the percent positivity data of NP testing (Miller et al., 2020). Both tests were assumed to have a specificity of 100%. We considered time delays of 24-48 hours in turnaround time from sampling to results, during which staff continued their shifts. Staff isolated for a period of 14 days following a positive result, during which they were excluded from interactions in the LTCF, and substitutes were included.

#### Vaccination

Considering LTCFs as one of the priority groups for COVID-19 vaccination, we implemented a two-dose vaccine strategy, with coverages of 70% for staff and 90% for residents (Government of Canada, 2021). We reviewed and extracted estimates of efficacy for Moderna vaccines following 2 doses, administered 28 days apart (Baden et al., 2020; U.S. Food and Drug Administration, 2020). We implemented a 14-day interval after the first dose of vaccine during which there was no statistically significant difference between the protection in the vaccinated and unvaccinated cohorts (Appendix, Table A2). Clinical trials indicated that vaccines were highly effective in individuals with predisposing medical conditions that put them at risk for severe Covid-19 (Baden et al., 2020). As sensitivity analyses, we also considered a scenario in which vaccine efficacies were reduced by a factor of *q* in vaccinated residents, where *q* was sampled uniformly from the 10%-50% range based on observed reductions in influenza vaccine effectiveness among frail and comorbid individuals (Andrew et al., 2017; Dhakal and Klein, 2019). The results of the sensitivity analysis are presented in Figure A3 of the Appendix.

### Model implementation and calibration

The model was implemented in Julia language using parameter estimates in Table 1. To determine the transmission probability, we calibrated the model to the cumulative incidence data reported for LTCFs in Ontario, Canada, from January to June 2020 (Public Health Ontario, 2020). For this calibration, we considered case isolation and hospitalization of infected residents, screening of staff for symptomatic illness, and mask-wearing by all staff as measures implemented during initial outbreaks of COVID-19. In the scenario with vaccination, we assumed that the first dose of vaccine was given two weeks before the introduction of the first infection into the LTCF, which resulted in partial effectiveness of vaccination when the outbreak simulations began. The second dose was offered two weeks after the start of simulations. Simulations were seeded with one infected individual among staff with a time-step of 1 hour. The computational model is available at https://github.com/thomasvilches/LTCF-covid.

## Results

In the baseline scenario of model calibration, the attack rate was 24.9% (95% CrI: 24.4 − 25.4) among residents, and 10.6% (95% CrI: 10.3 − 10.8) among staff over a 200-day time horizon. Hospitalization and deaths among residents were projected to be 21.1 (95% CrI: 20.6 − 21.8) and 52.2 (95% CrI: 51.0 − 53.4) per 1000 population, respectively, over the same time horizon. The corresponding rates for staff were 2.8 (95% CrI: 2.6 − 3.0) and 0.35 (95% CrI: 0.28 − 0.43) per 1000 population.

### Routine testing of staff

Compared to baseline measures only, the addition of weekly routine testing of staff reduced the attack rate among residents by 25.9% (95% CrI: 23.3% − 28.3%) using saliva sampling with a 2-day turnaround. The highest reduction was 42.1% (95% CrI: 40.3% − 43.9%) using weekly NP sampling with a 1-day turnaround time (Figure 1C, Table 2). For each scenario, the observed reductions of attack rates among staff were similar to those among residents (Table 2).

**Table 2.** Mean and 95% credible intervals for the reduction of cumulative infections, hospitalizations, and deaths among residents achieved by additional measures of 7-day routine testing of staff, and vaccination of staff and residents as compared with baseline measures alone, over a 200-day time horizon.

| Measure |  | Mean relative reduction (%) and 95% CrI |  |  |
| --- | --- | --- | --- | --- |
| 7-day routine testing |  | Infection | Hospitalization | Death |
| NP sampling | 1-day turnaround | 42.1 (40.3, 43.9) | 42.4 (39.7, 45.2) | 43.2 (41.3, 45.4) |
|  | 2-day turnaround | 30.3 (28.4, 32.3) | 29.9 (26.6, 33.0) | 31.0 (28.9, 33.2) |
| Saliva sampling | 1-day turnaround | 34.3 (32.2, 36.2) | 34.4 (31.1, 37.1) | 34.6 (32.4, 36.9) |
|  | 2-day turnaround | 25.9 (23.3, 28.3) | 23.9 (20.4, 27.1) | 26.1 (23.5, 28.6) |
| Vaccination |  | 88.5 (88.1, 88.9) | 95.9 (95.4, 96.3) | 95.8 (95.5, 96.1) |

Weekly routine testing of staff led to a similar reduction of hospitalizations among residents, ranging from 23.9% (95% CrI: 20.4% − 27.1%) to 42.4% (95% CrI: 39.7% − 45.2%) with a 2-day turnaround for saliva testing and a 1-day turnaround for NP testing, respectively (Figure 1C, Table 2). The corresponding reductions in deaths among residents ranged from 26.1% (95% CrI: 23.5% − 28.6%) to 43.2% (95% CrI: 41.3% − 45.4%). We observed similar reductions of hospitalizations and deaths among staff (Table 3).

**Table 3.** Mean and 95% credible intervals for the reduction of cumulative infections, hospitalizations, and deaths among staff achieved by additional measures of 7-day routine testing of staff, and vaccination of staff and residents as compared with baseline measures alone, over a 200-day time horizon.

| Measure |  | Mean relative reduction (%) and 95% CrI |  |  |
| --- | --- | --- | --- | --- |
| 7-day routine testing |  | Infection | Hospitalization | Death |
| NP sampling | 1-day turnaround | 41.5 (39.3, 43.8) | 38.6 (31.9, 45.0) | 45.4 (24.4, 59.9) |
|  | 2-day turnaround | 29.5 (27.2, 31.7) | 25.3 (17.2, 31.9) | 24.4 (0.01, 43.3) |
| Saliva sampling | 1-day turnaround | 33.4 (31.0, 35.5) | 28.4 (23.0, 36.8) | 28.8 (0.1, 46.4) |
|  | 2-day turnaround | 25.3 (22.6, 28.1) | 26.2 (16.9, 33.8) | 20.5 (-0.04, 37.7) |
| Vaccination |  | 88.9 (88.4, 89.4) | 87.8 (84.3, 90.6) | 89.7 (83.5, 95.9) |

### Vaccination of residents and staff

When vaccination was implemented, the incidence among residents sharply declined after 4 weeks (Figure 1D), as compared with baseline measures alone. The attack rate was reduced by 88.5% (95% CrI: 88.1% − 88.9%) among residents and 88.9% (95% CrI: 88.4% − 89.4%) among staff over a 200-day time horizon (Tables 2, 3), significantly higher than reductions achieved in any scenario of routine testing in both populations (Wilcoxon rank-sum test, p-values < 0.001). Vaccination reduced hospitalizations and deaths among residents by 95.9% (95% CrI: 95.4% − 96.3%) and 95.8% (95% CrI: 95.5% − 96.1%); the corresponding reductions in staff were 87.8% (95% CrI: 84.3% − 90.6%) and 89.7% (95% CrI: 83.5% −95.9%) (Figure 1F, Tables 2, 3).

## Discussion

The COVID-19 pandemic has disproportionately affected geriatric populations and residents of LTCFs with devastating outcomes (Barnett and Grabowski, 2020; Bell et al., 2020; Burton et al., 2020; Canadian Institute for Health Information, 2020; ECDC Public Health Emergency Team et al., 2020). The incidence rate ratio for COVID-19 related deaths among LTCF residents in Ontario, Canada has been estimated to be 13 times higher than that among adults older than 69 years in community living residences (Fisman et al., 2020), with case fatality rates exceeding 27% (Brown et al., 2020) during the first pandemic wave. In the absence of vaccination, the control of COVID-19 outbreaks in these vulnerable settings has proved to be challenging largely due to staff shortages and frequent staff turnover, low staff-to-resident ratios, crowded settings without room to implement physical distancing, insufficient training, as well as silent transmission of disease. Furthermore, reliance on symptom-based screening can lead to the introduction of COVID-19 in LTCFs by asymptomatic, or even mild symptomatic staff which may result in widespread transmission.

Our evaluation of multi-pronged strategies in LTCFs indicate that augmenting non-pharmaceutical measures with routine testing of staff can reduce the rates of silent transmission and would decrease infections and adverse clinical outcomes considerably among residents. We found that with a 2-day or longer time-delay from sample collection to results, the impact of this measure on reducing hospitalizations and deaths among residents was decreased by at least 12% and 8% compared to a 1-day turnaround time in NP and saliva testing, respectively, over a 200-day time horizon (Figure 1C, Table 2). We observed similar effects of a time-delay in routine testing of staff on cumulative infections among residents (Figure 1B, Table 2). Vaccination would significantly (2-4 times more than routine testing of staff) reduce infection and severe outcomes among both residents and staff. While vaccines reduce the incidence of infection and may decrease transmission, the practice of other measures (i.e., mask-wearing, social distancing, hand hygiene) will still be needed for some time until community transmission is controlled. Encouragingly, in the province of Ontario, these non-pharmaceutical measures have remained in place despite vaccination of the majority of residents and staff.

### Strengths and limitations

A key strength of our dynamic transmission model is the integration of several interventions implemented in LTCFs (i.e., mask wearing, symptom-based screening, staff cohorting, isolation, routine testing, and vaccination) and evaluation of their effectiveness in controlling outbreaks in the context of the COVID-19 pandemic. We also utilized interaction data collected using sociometric devices in a real setting (Champredon et al., 2018; Najafi et al., 2017) to parameterize the contact patterns in the model, as well as observed contacts between individuals during LTCF outbreaks in Ontario, Canada. However, our results should be interpreted within the study assumptions and limitations. For the model structure, staff-to-resident ratio, and population interactions, we relied on existing data and correspondence with LTCFs affected by COVID-19 in Ontario, Canada. We did not include visitation by community members in the model during the outbreak, which may be allowed with specific guidelines for visitors. We also did not include other modes of disease transmission such as aerosolization of the virus without adequate ventilation. For evaluation of routine testing, we assumed a 100% compliance rate among staff. While this may be a reasonable assumption for non-invasive, self-administered saliva testing, compliance will likely be affected by practical challenges of relatively invasive NP testing. We assumed that staff cohorting can be effectively and sustainably implemented as described; however, staff shortages and the use of overtime to replace staff may affect the effectiveness of this strategy.

## Conclusions

Our study highlights the importance of multifaceted strategies, including non-pharmaceutical measures and vaccination, for protecting vulnerable residents in LTCFs. Without population-level control of disease, the risk of infection and silent transmission by staff or visitors in these settings cannot be discounted even with a highly efficacious vaccine.

## Data Availability

The computational model was implemented in Julia language and is available at: https://github.com/thomasvilches/LTCF-covid.

## Author Contributions

SMM, LEC, APG, and JML were responsible for conception and design. TNV and SN developed the model and performed simulations. SMM and KZ analyzed the data and drafted the results. LJK and PS parameterized the model and contributed to collection of data and resources. All authors contributed to writing, interpreting the results, and revising the manuscript for accuracy of its content. SMM is the guarantor.

## Additional Contributions

The authors thank the management teams of LTCFs in Ontario, Canada for helpful conversations that informed our analysis, strategies, and assumptions.

