## Supplemental FIle for "Multifaceted strategies for the control of COVID-19 outbreaks in long-term care facilities in Ontario, Canada"

This appendix provides further details of the model structure, its parameterization, and the results of vaccination scenario with reduced efficacy in residents.

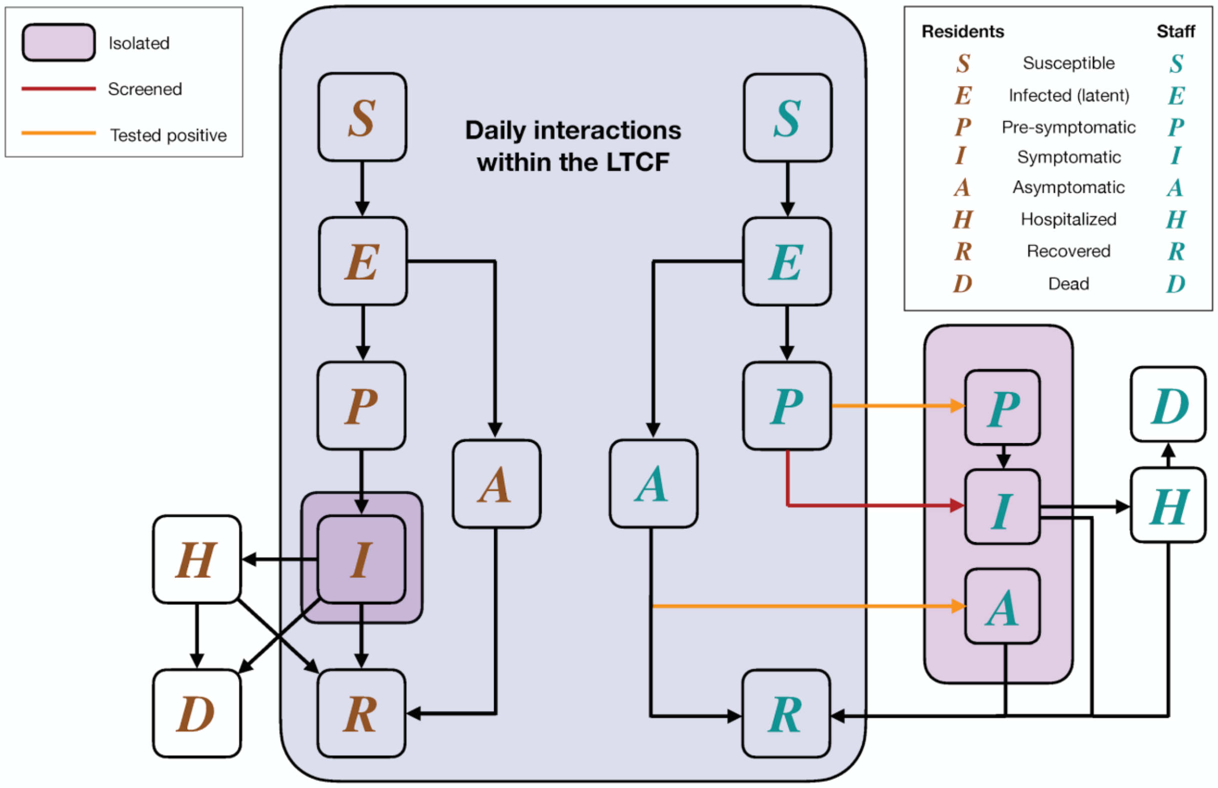


**Figure A1.** Schematic model diagram for disease dynamics and interventions.

### *Daily number of contacts*

The staff-to-resident ratio was informed through correspondence with the management teams of 10 Ontario LTCFs with COVID-19 outbreaks. The model includes one personal support worker per nine residents during day and evening shifts, and one per 22 residents in the night shift. The ratio of nurses to residents was one to 32 during day and evening shifts and one to 64 in the night shift. The ratio of dietary and housekeeping staff to residents was one to 32 in all shifts.

The daily number of contacts between residents was sampled from a previously inferred distribution with a mean of 6.8 contacts per day per resident [1,2]. The number of contacts that a resident had with a personal support worker was one to two per shift. Each resident had one contact with a nurse and one contact with a dietary staff per shift. Residents were assumed to have one contact daily with housekeeping staff. Contacts are summarized in Table A1.

**Table A1.** Mean number of daily contacts among a single agent and a group of agents.

<table>
<tr><th rowspan="2">Single Agent</th><th colspan="7">Group of Agents</th></tr>
<tr><td colspan="3">Resident</td><td>PSW</td><td>Nurse</td><td>DS</td><td>HS</td></tr>
<tr><td>Resident</td><td colspan="3">6.8 (SD: 4.75)</td><td>3-6</td><td>3</td><td>3</td><td>1</td></tr>
<tr><td>Personal support worker (PSW)</td><td>Day<br>8-18</td><td>Evening<br>8-18</td><td>Night<br>20</td><td colspan="4">2-4</td></tr>
<tr><td>Nurse</td><td>30</td><td>30</td><td>60</td><td colspan="4">2-4</td></tr>
<tr><td>Dietary staff (DS)</td><td>30</td><td>30</td><td>30</td><td colspan="4">2-4</td></tr>
<tr><td>Housekeeping staff (HS)</td><td>15</td><td>15</td><td>0</td><td colspan="4">2-4</td></tr>
</table>

**Table A2.** Estimated efficacies of Moderna vaccines with associated timelines following each dose.

| **Vaccine efficacy** | **Week after the first dose** | | **Week after the second dose** | |
|---|---|---|---|---|
| Moderna | 1-2 | 3-4 | 1-2 | >2 |
| Against infection | None | 61% (31% – 79%) | 61% (31% – 79%) | 93.5% (85.2% - 97.2%) |

| | | | | |
|---|---|---|---|---|
| Against symptomatic disease | None | 92.1% (68.8% - 99.1%) | 92.1% (68.8% - 99.1%) | 94.1% (89.3% - 96.8%) |
| Against severe disease | None | 92.1% (68.8% - 99.1%) | 92.1% (68.8% - 99.1%) | 100% |

***Routine testing of staff***

We implemented the temporal diagnostic sensitivity of NP and saliva testing derived from our previous work [3] by fitting a sensitivity function to data reported for percent positivity of COVID-19 patients. Figure A2 shows the model outputs for the proportion of silent infections (i.e., pre-symptomatic or asymptomatic infection) detected among staff by a 7-day frequency of NP and saliva testing. Case identification is affected by both sensitivity of the test and the time from sample collection to the laboratory results, which affects the dynamics of infection in the LTCF.

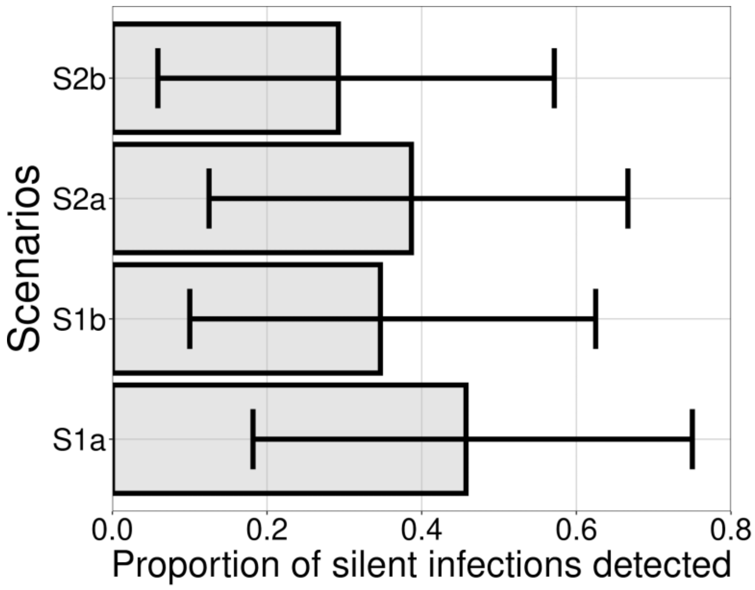


**Figure A2.** Proportion of silent infections that are detected in routine testing of staff. Scenarios correspond to NP testing with 1-day (S1a) and 2-day (S1b) turnaround times, and saliva testing with 1-day (S2a) and 2-day (S2b) turnaround times.

***Results with reduced vaccine efficacy for residents***

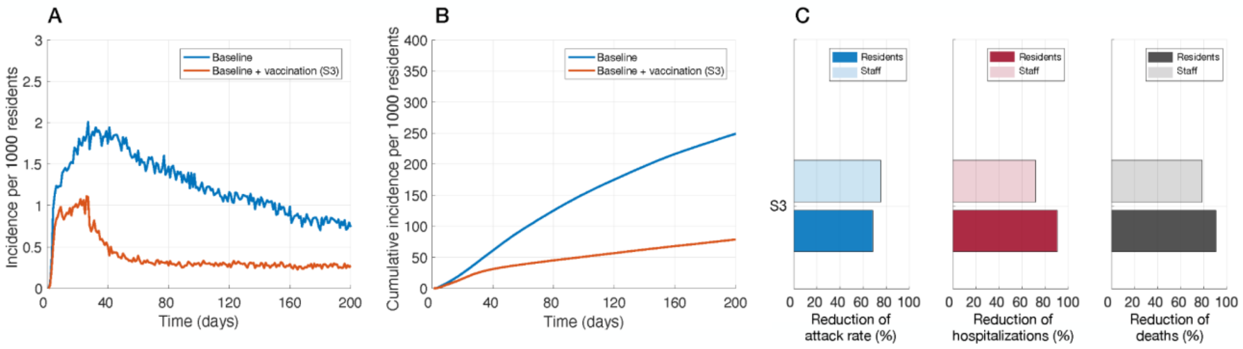


**Figure A3**. Incidence of infection per 1000 residents (A), cumulative infections per 1000 residents (B), and relative reduction of cumulative infections (attack rate), hospitalizations, and deaths with vaccination of residents and staff (C), over a 200 time period of simulations.

**Table A3.** Mean and 95% credible intervals for the reduction of cumulative infections, hospitalizations, and deaths among residents and staff attributed to vaccination of staff and residents as compared with baseline measures alone, over a 200-day time horizon.

| **Measure** | **Mean relative reduction (%) and 95% CrI** | | |
|---|---|---|---|
| Vaccination | Infection | Hospitalization | Death |
| Residents | 68.4 (67.5, 69.4) | 90.4 (89.7, 91.0) | 90.7 (90.2, 91.1) |
| Staff | 75.6 (74.6, 76.5) | 71.6 (67.1, 75.8) | 79.2 (67.0, 88.7) |
